# The Impact of Spinal Manipulation on Lumbar Proprioception and its Link to Pain Relief: A Randomized Controlled Trial

**DOI:** 10.1101/2024.11.10.24316819

**Authors:** Nyirö Luana, Dörig Monika, Suter Magdalena, Connolly Lukas, Vogel Noemi, Carla Stadler, Giovanna John-Cecere, Schweinhardt Petra, Meier Michael Lukas

## Abstract

Manual therapy, such as spinal manipulation (SM), is commonly used to treat non-specific chronic low back pain (CLBP), although its mechanisms remain poorly understood. It has been hypothesized that the mechanical forces applied during spinal manipulation (SM) influence proprioceptive function, which is often impaired in patients with CLBP. This study aimed to investigate the effect of a single SM intervention on lumbar proprioceptive function and its potential relationship with analgesic effects in patients with CLBP. In a single-blind randomized controlled trial, data from 142 adults with or without CLBP were analyzed after random assignment to receive lumbar spinal manipulation (LMANIP), lumbar mobilization (LMOB), or no intervention (NI). The primary outcome was the proprioceptive weighting (PW) ratio, which reflects the central nervous system’s preferred source of proprioceptive input for balance control, specifically from the lumbar and ankle muscles. PW ratios were assessed immediately before and after intervention by analyzing postural sway changes during vibrotactile stimulation (60 Hz). PW changed in both healthy participants and patients after the intervention, with a significantly stronger lumbar-steered PW following LMANIP compared to NI (β = -0.047, t(422) = -2.71, p = 0.007) and LMOB (β = -0.039, t(422) = - 2.17, p = 0.030). Moreover, LMANIP was particularly effective in reducing pain in patients with stronger lumbar-steered PW before intervention (p < 0.017). These findings suggest that a single SM session enhances proprioceptive input from the lumbar muscles and that the strength of the analgesic effect is associated with the baseline PW status.

## 1. Introduction

Low back pain (LBP) is one of the most prevalent health conditions, affecting approximately 80 % of individuals at some point in their lives [22,33,54], and is a leading global health issue in terms of years lived with disability [25,50]. In the majority of LBP cases, there is no pathoanatomical cause identifiable [54], leading to a diagnosis of ’non-specific LBP’ [33]. Many patients do not fully recover and experience recurrent episodes [32]. Consequently, chronic LBP (CLBP), defined as pain lasting ≥ 12 weeks, causes considerable personal, economic, and social costs [10,25]. Spinal manipulation (SM) therapy is a frequently used and guideline-recommended non-pharmacological intervention for both acute [38,43,59] and chronic LBP [37,56]. Although SM can relieve pain and improve function [43,60], the precise mechanisms underlying SM’s beneficial effects remain poorly understood. Several mechanisms have been proposed, including biomechanical [16,23,57] and neurophysiological effects observed at the spinal [39,58] and supraspinal levels [11,19], with an unclear relationship to potential analgesic effects [58]. In recent years, evidence for neurophysiological effects has increased, as several studies have reported changes in somatosensory processing and muscle reflex responses following SM [19]. A neurophysiological mechanism potentially associated with the clinical benefits of SM may involve its ability to modulate the firing rate of muscle spindles [40,52], the principal receptors for proprioception, which is often impaired in patients with LBP [51]. Proprioceptors can play a direct pro-nociceptive role in chronic pain states [31], and proprioceptive dysfunction has been suggested to indirectly provoke or maintain LBP through maladaptive/increased loading on spinal tissues, due to impaired paraspinal sensory feedback that negatively affects trunk motor control [26,35,36]. Proprioceptive weighting (PW) is a crucial function of the human proprioceptive system, which is the capability of the central nervous system to selectively prioritize the most reliable proprioceptive inputs from key body stabilizers, such as the ankle and lumbar muscles, which are essential for maintaining posture and effective motor control [4,6]. PW can be assessed in humans using balance control measures (based on recordings from a force plate) in combination with vibrotactile stimulation at 60 Hz, which mostly activates primary afferents in the muscle spindles [5,27,49]. Impaired PW capacity has been demonstrated in patients with LBP [4,5,27], characterized by over-reliance on proprioceptive input from the ankle muscles [5]. In addition, ankle-steered PW in healthy subjects has been associated with an increased risk of developing LBP [7]. Interestingly, SM-like loads applied to the spinous process of the L6 vertebra in anesthetized cats increased the discharge frequency of paraspinal muscle spindles [40]. This suggests that SM in humans may modulate PW, which could be significant in terms of both treatment and prevention of LBP. However, evidence of such a neurophysiological effect and its association with clinical outcomes of SM in patients with LBP is lacking. This study aimed to investigate whether a single SM intervention has an effect on PW. We hypothesized that SM enhances proprioceptive input from the lumbar muscles, potentially shifting PW towards the lumbar muscles and away from the ankle. In addition, we explored whether PW is associated with the analgesic response to SM.

## 2. Methods

### 2.1 Study Design

This study was designed as a single-blind randomized controlled trial of lumbar spinal manipulation (LMANIP) vs. lumbar spinal mobilization (LMOB) vs. no intervention (NI). Initially, the study aimed to include only healthy participants and featured an additional treatment arm for thoracic manipulation to examine location-specific effects. However, to increase statistical power, the thoracic intervention arm was removed before recruitment began. In addition, the study was expanded to include patients with LBP. While this addition enriched the clinical relevance of the study, a dedicated sample size calculation for the patient group was not conducted (for more information see 2.8 and limitations). Data were collected at the Balgrist University Hospital, Zurich, Switzerland. The trial was preregistered at ClinicalTrials.gov (NCT 04869514) and approved by the Swiss local ethics board ‘Kantonale Ethikkommission Zürich, KEK’ (Nr. 2021-01331). All study procedures adhered to the Declaration of Helsinki and current legislation pertaining to the management of personal data. The results are reported in accordance with the Consolidated Standards for Reporting Trials (CONSORT) statement [46]. This study consisted of a single visit (Figure 1).

**Figure 1.**
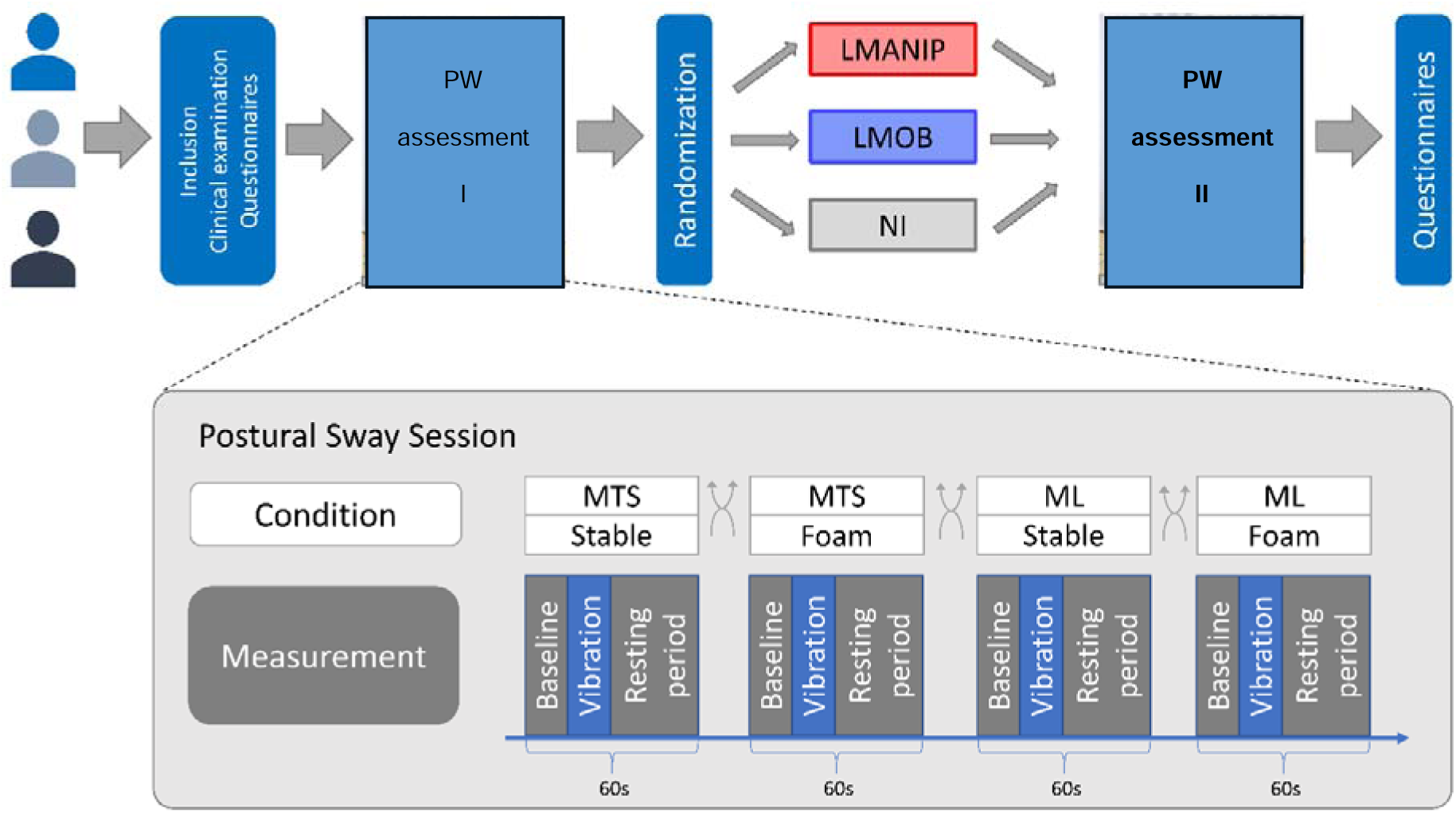
Study Design. LMANIP, lumbar spinal manipulation; LMOB, lumbar spinal mobilization; NI, no intervention; MTS, M. triceps surae; ML, M. longissimus.

### 2.2 Participants

Healthy individuals and patients with non-specific CLBP were recruited for this study. Participants had to meet the following eligibility criteria: age between 18 and 50 years, and no history of vestibular disorders. For healthy participants, additional inclusion criteria included: no episode of musculoskeletal pain in the past 3 months, and no history of chronic pain. Specific inclusion criteria for CLBP patients were a history of LBP for more than 3 months, clinically not attributable to "red flags" (e.g. infection, trauma, fractures, or inflammatory spondyloarthropathies), no episode of musculoskeletal pain other than LBP in the past 3 months, and no history of chronic pain other than CLBP. Participants were excluded if they met any of the following criteria: excessive consumption of alcohol (exceeding recommended limits within a single day, i.e. more than 4 standard drinks for men and more than 3 for women), drugs, or analgesics within the last 24 hours; pregnancy or breastfeeding; previous foot, ankle, or spine surgery; recent chiropractic or other manual treatments within the last two weeks; neuromuscular disorders potentially impacting gait and posture; injuries to the motor system resulting in permanent deformities; a body mass index (BMI) below 16 kg/m² or above 30 kg/m²; contraindications to SM interventions; or inability to tolerate SM. For patients with CLBP, additional exclusion criteria included any facet joint, epidural, or periradicular injections within the past six months.

### 2.3 Recruitment

Participants were recruited via advertisements through social media, online platforms, and mailing lists of the University of Zurich and among the staff of the Balgrist University Hospital (by advertisements and oral communication), with particular caution that the recruited persons were not in a relationship of dependence to any of the study staff and were not familiar with the concepts of PW. Participants were screened via an interview and clinical examination, performed by experienced study staff with a degree in physiotherapy and more than three years of clinical experience, during the study visit. The clinical examination included a neurological examination and palpation of the most painful spinal segment in all patients. Participants who met all the eligibility criteria were included in the study.

### 2.4 Vibrotactile stimulations and movement illusions

All participants underwent a postural sway session before and immediately after the intervention. The participants stood on a force plate (Kistler®, Winterthur, Switzerland; type 9260AA6, sampling rate 1000 Hz) with their feet shoulder-width apart and arms hanging loosely at the side. Vision was occluded using non-transparent goggles, and participants wore noise-cancelling headphones. Each PW assessment session included four conditions of bilateral vibrotactile stimulation (Figure 1). Each condition (total duration of 60 s) included a baseline measurement (15 s), vibratory stimulation (15 s), and rest period (30 s). Stimulation was applied to the muscle-tendon junction of the calf muscles (M. triceps surae (MTS)) and paraspinal muscles (M. longissimus (ML)) at the lumbar level while standing on stable and unstable support surfaces (using a foam pad, fixed on the force plate). This approach is commonly used to assess how different support surfaces influence proprioceptive strategies [4,27]. Vibrotactile stimulation was applied using an electrical vibration unit (custom-made, Hasselt University, Diepenbeek, Belgium), enabling targeted vibrations with a frequency of 60 Hz. The stimulation sites were chosen to affect skeletal muscles that have been shown to be important in providing proprioceptive information for balance [4]. The order of the stimulations (MTS/ML) and conditions (stable/foam) was randomized using a random number sequence generator in R, generating a predetermined allocation sequence. The research assistants consecutively allocated the participants to the allocation sequence as they entered the study. At the end of the second PW assessment, participants were asked whether they experienced movement illusions during MTS and/or ML vibrotactile stimulation, because these perceptions are considered an indicator of successful placement of the vibrator and stimulation of muscle spindles [49].

### 2.5 Interventions

Participants were randomly assigned to one of the interventions: 1) LMANIP, 2) LMOB, or 3) NI. LMANIP involved high-velocity low-amplitude (HVLA) SM at the L4/L5 motion segment applied in side-posture bilaterally, with a hypothenar contact applied to the right or left L5 mammillary process to induce gapping of the ipsilateral L4/5 articulation (resisted positioning, with contact applied to the inferior vertebra to induce gapping in the joint superior to the contact), and the treatment force directed posterior to anterior, in the facet joint-plane [2,18,24].

LMOB involved the same positioning and manual contact as described for the LMANIP intervention, but instead of an HVLA thrust, a Grade III mobilization, described by Maitland as a large-amplitude movement performed up to the limit of the range, was applied with a duration of 30 s and frequency of 1 Hz on each side [34]. The NI group was instructed to rest in a side-lying position for 30 s on each side, and no manual contact was applied. The order in which the intervention was applied to the left or right side was pseudo-randomized for all participants. All interventions were performed by an experienced clinician (more than five years of clinical experience).

### 2.6 Randomization and masking

The treatment allocation (LMANIP, LMOB, NI; 1:1:1 allocation ratio) was placed in a sealed opaque envelope, each envelope identical in appearance and containing a card indicating the assigned treatment. The sealed envelopes were thoroughly mixed to ensure randomness and to prevent potential bias. After completion of the baseline questionnaires, the participants were instructed to draw an envelope from a box containing the envelopes. To ensure that the treatment assignment remained concealed until the intervention, the participant handed over the envelope to the treating clinician who opened the envelope in the presence of the participant to ensure transparency.

The research staff conducting the clinical examinations and data collection were blinded to the intervention. The clinician delivering the intervention was not involved in any data collection and was blinded to whether the participant was a healthy subject or patient. Participants were blinded to the study hypotheses. At the end of the study visit, an exit interview was conducted in which the study staff had to state to which of the three interventions they thought that the participants had been assigned, and the clinician had to state whether she thought the participant was a healthy control or a patient. The investigator conducting the statistical analyses was blinded to the treatment identity/group allocation.

### 2.7 Outcomes

The primary outcome measure was the PW ratio, calculated based on the anterior-posterior displacement of the center of pressure (COP) during vibrotactile stimulation of the MTS and ML. COP displacements (in mm) were derived from changes in the Y-coordinate of the COP and calculated from raw force plate data at a temporal resolution of 1000 Hz using the formula COP = Mx/Fz [4,5] (Figure 2). COP excursions were analyzed exclusively in the sagittal plane to enhance sensitivity, as COP displacement occurs anteriorly following lumbar muscle vibration and posteriorly following triceps surae muscle vibration [4,5]. The COP displacements were calculated as the absolute value of the difference between the mean COP position in the y-direction during a 15 s baseline period (vibration off) and during a 15 s period of muscle vibration (vibration on). Positive values indicate anterior COP displacement, while negative values indicate posterior COP displacement.

**Figure 2.**
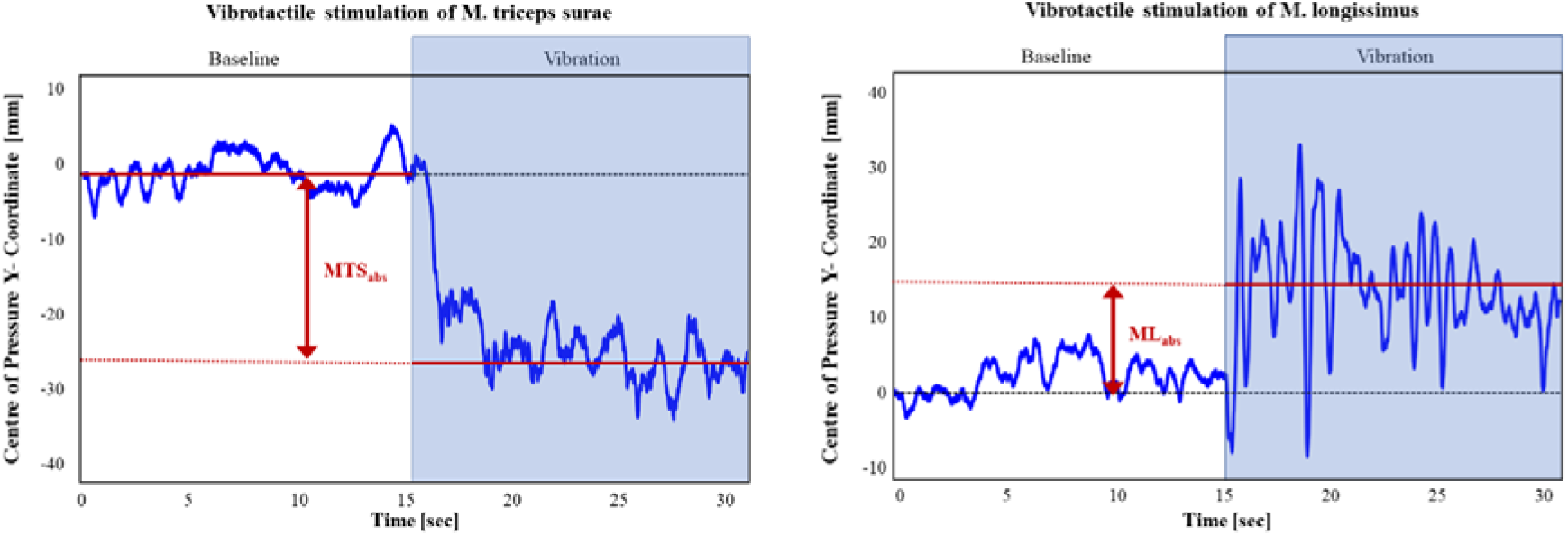
Postural sway paths. Illustrative example of single-subject data showing changes in the Y-coordinate of the center of pressure (COP). The measure used was the absolute displacement of the mean COP in millimeters (mm, red line), reflecting the anterior-posterior displacement. MTSabs represents the absolute displacement of the mean COP [mm] during M. triceps surae vibration, whereas MLabs represents the absolute displacement of the mean COP [mm] during M. longissimus vibration.

PW ratios were calculated using the following formula:

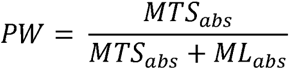

similar to the method reported by Brumagne et al. [5] (MTS_abs_ = absolute anterior-posterior displacement of the mean COP [mm] during MTS vibration, ML_abs_ = absolute anterior-posterior displacement [mm] during ML vibration). A ratio closer to ”1” corresponds to more reliance on ankle proprioception, while a ratio closer to ”0” indicates more reliance on lumbar proprioception.

Secondary outcomes were the scores from (1) the Tampa Scale of Kinesiophobia (TSK), (2) Spielberger’s State and Trait Anxiety Inventory (STAI), (3) Baecke Habitual Physical Activity Questionnaire (BQHPA), (4) Fremantle-back awareness questionnaire (FreBAQ), (5) numeric rating of the participant’s belief that a manual intervention will relieve pain (healthy participants were asked to imagine having pain), and (6) numeric rating (before intervention) of the participant’s belief that a manual intervention will affect balance control. In patients, additional outcomes included (6) numeric pain rating scale (NPRS) of clinical pain before and after the intervention, (7) Oswestry disability index (ODI), and (8) PainDetect questionnaire. The TSK questionnaire contains statements focusing on fear of physical activity and is used to assess the fear of movement [45,53]. The STAI is a common questionnaire that measures state and trait anxiety levels [47], BHQA measures self-reported physical activity [1], and the FreBAQ measures back-specific body-perception [55]. The ODI is commonly used to assess disability, and painDETECT is used to assess current pain, strongest, and average pain intensity in the previous 4 weeks [12,14].

### 2.8 Statistical Analysis

Assuming a medium to large effect size (Cohen’s d = 0.7) based on F tests (statistical tests for ANOVA: fixed effects, main effects, and interactions), alpha = 0.05, and power 0.8, a total sample size of 84 subjects was required. In anticipation of study dropouts and a potentially smaller effect size, we determined a total sample size of 120 healthy participants (40 participants per intervention group). A dedicated sample size calculation for the patient group was not conducted. Statistical analyses were performed using R 4.4.2 for Windows with the software package RStudio version 2024.04.2 (Boston, MA, 2022), and the open-source software Jamovi (The jamovi project (2021), version 2.5.3, www.jamovi.org). Statistical significance was set at p < 0.05.

Normal distribution of raw data or of the residuals of the models, depending on the statistical test used, was evaluated by assessing z-scores of skew and kurtosis, which were required to be smaller than 2, and by visual assessment of histograms and Q–Q plots. Participants’ baseline characteristics are described as mean values and SDs for normally distributed continuous data or medians and ranges for continuous data with skewed distributions. Categorical variables were analyzed using frequency analysis and are presented as counts and percentages.

#### 2.8.1 Mixed model with PW ratio (primary outcome) as dependent variable

A linear mixed model was fitted to assess the effects of intervention and other predictors on the PW ratio. The model included fixed effects for "sex” (male/female), "age", "bmi", "intervention" (LMANIP/LMOB/NI), "surface" (stable/foam), “group" (healthy control/patient), "timepoint" (before intervention/after intervention), and their interaction ("timepoint:intervention"). Interaction effects of "age:sex", "intervention:surface", "intervention:group", "group:surface" and "group:timepoint" were first included in the model and removed afterwards because they were not significant to obtain an estimation of the main effects.

#### 2.8.2 Mixed model with pain intensity (NPRS score) as dependent variable

A linear mixed model was fitted using data from the patient group, with pain intensity (NPRS scores before and immediately after intervention) as the outcome variable, 1) to test for potential differential effects of intervention between before-/after pain ratings, and 2) explore whether baseline PW is a possible moderator of these effects. The following fixed effects were included: "sex" (male/female), "age", "bmi", "timepoint" (before/after intervention), "intervention" (LMANIP/LMOB/NI) and "baseline PW ratio" (mean of stable/foam surfaces, as these were significantly correlated (Spearman’s Rho = 0.432, p < 0.001), reducing redundancy in predictors and simplifying the model). Interaction terms incorporated in the model were "timepoint:intervention" and "timepoint:intervention:baseline PW ratio". Interaction effects of "age:sex", "intervention:surface", "intervention:group", "group:surface", and "group:timepoint" were first included in the model and removed afterwards.

For all models, a random effect "participant" was included to account for variability between subjects, with no specific assumption regarding the pattern of correlation between repeated measures. In the presence of a significant main or interaction effect, pairwise comparisons of the respective parameter estimates (fixed coefficients) were reported. To ensure that the results of the models were not driven by outliers, an assessment was conducted for each model to identify influential cases, defined as outliers that significantly impact the model’s outcome [13]. This process involved two main steps: 1) identification of data points that were outliers within the model, defined as having a z-score of the residual larger than 2 and 2) determining which of these outliers had substantial influence on the model, as indicated by Cook’s distance exceeding the threshold of 4/(n-k-1), where n is the number of data points and k is the number of predictors. To verify whether these influential data points affected the model’s conclusions, the model was re-run excluding the identified data points. As the exclusion of data points did not change the statistical inference of the model, the results of the full models were reported.

## 3. Results

### 3.1 Recruitment and subject characteristics

Recruitment started in May 2023, enrollment ended in April 2024, the first study visit was 11.05.2023, and the last study visit was completed on 08.04.2024. No additional healthy participants were scheduled to participate when the target of 120 healthy participants was reached. Those who were already scheduled for a visit were assessed for eligibility and were potentially included. In total, 254 participants were recruited for the study, of whom 206 were randomly assigned to the interventions (separately per group). 70 were allocated to the LMANIP intervention group, 66 to the LMOB intervention group, and 66 to the NI control group. Data from 64 individuals were excluded from the analysis: 4 due to inconsistent information regarding the presence or duration of pain and 60 due to the lack of movement illusions in at least one stimulation location (sections 3.2 and 4). A total of 142 participants (90 healthy participants and 52 patients) were included in the final analysis (Figure 3).

**Figure 3.**
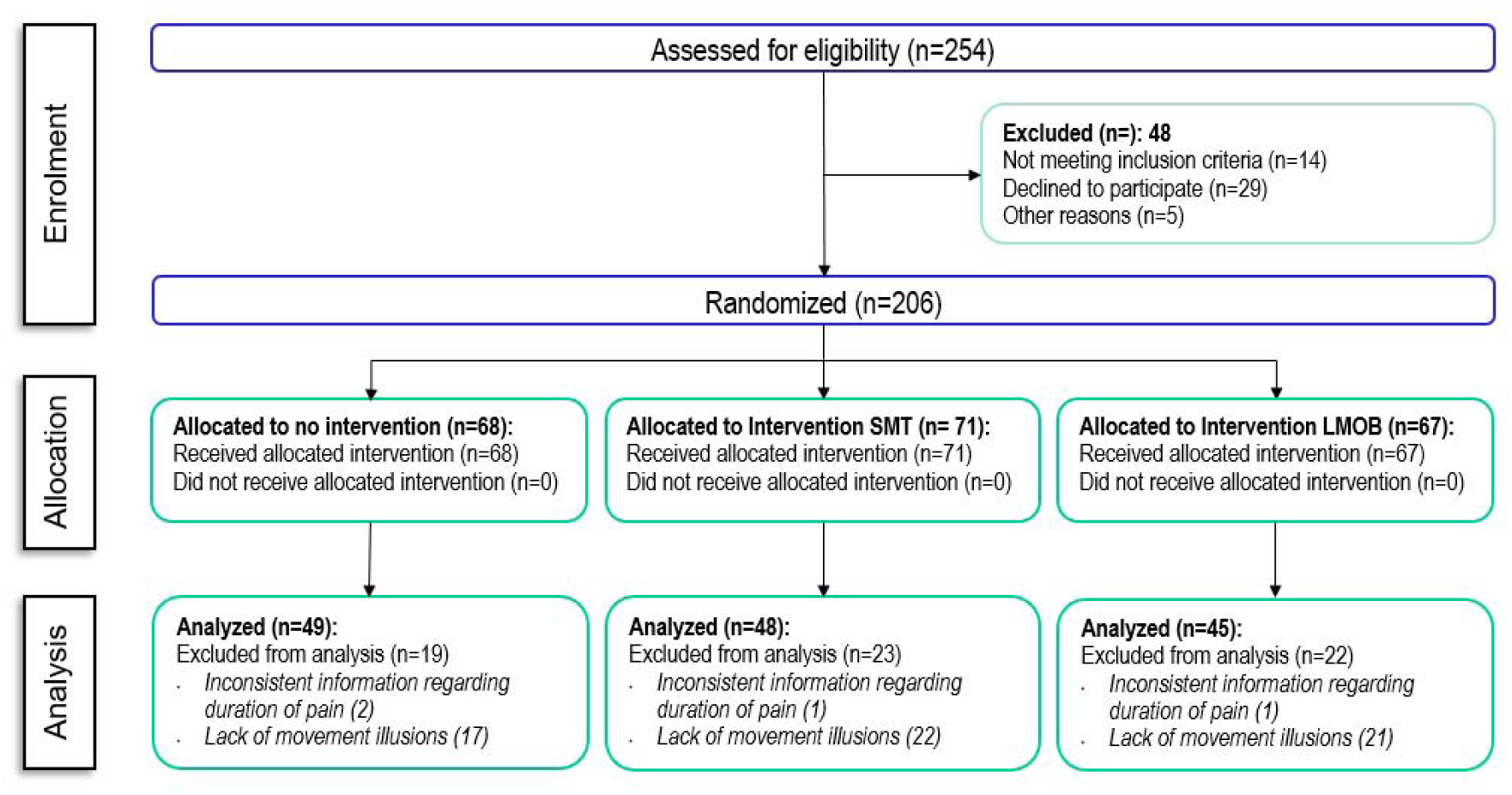
Flowchart of the study. NI, no intervention; LMANIP, spinal manipulation; LMOB, spinal mobilization.

### 3.2 Movement illusions

Sixty participants reported no movement illusions in at least one location (9 participants at both locations). Sixteen participants (13 healthy participants and 3 patients) reported no illusions during MTS stimulation, whereas 53 participants reported no illusions during ML stimulation (41 healthy participants and 12 patients). The presence or absence of movement illusions during ML stimulation significantly affected postural sway (supplementary material S2). These 60 “non-responders” were excluded from the final analysis because the absence of movement illusions indicates a failure of appropriate stimulation of the proprioceptive system [49] and can therefore distort the primary outcome (PW ratio).

### 3.3 Baseline characteristics

There were no significant differences in age or BMI across interventions (p’s > 0.06). Significantly fewer women were randomized to the LMANIP group (p=0.003). There were no significant differences in the expectation of balance (p = 0.769) or pain (p = 0.591) across the interventions. For more information regarding demographics, refer to Table 1.

**Table 1.** Baseline characteristics.

|  | Lumbar Manipulation |  | Lumbar Mobilization |  | No Intervention |  | p |
| --- | --- | --- | --- | --- | --- | --- | --- |
|  | Healthy Controls | Patients | Healthy Controls | Patients | Healthy Controls | Patients |  |
| n | 31 | 17 | 27 | 18 | 32 | 17 |  |
| Gender = Female (%) | 15 (48.4) | 7 (41.2) | 22 (81.5) | 13 (72.2) | 22 (68.8) | 13 (76.5) | 0.003 |
| Age (years) | 25.00 [23.50, 28.50] | 29.00 [24.00, 32.00] | 25.00 [24.00, 30.00] | 26.00 [24.00, 31.50] | 23.00 [21.80, 26.00] | 26.00 [23.00, 32.00] | 0.067 |
| Weight (kg) | 66.50 [58.50, 72.40] | 75.4 [60.40, 85.30] | 65.30 [55.60, 72.50] | 71.70 [55.70, 79.00] | 60.09 [56.30, 65.00] | 68.30 [58.9, 72.50] | 0.027 |
| Height (cm) | 175.00 [170.00, 180.00] | 176.00 [170.00, 186.00] | 169.00 [162.00, 177.00] | 171.00 [164.00, 176.00] | 169.00 [164.00, 173.00] | 167.00 [164.00, 171.00] | 0.001 |
| BMI (kg/m<sup>2</sup>) | 21.40 [20.40, 23.80] | 22.60 [20.30, 24.10] | 22.9 [21.00, 24.20] | 23.50 [20.20, 26.60] | 21.3 [20.30, 22.80] | 23.50 [22.40, 25.40] | 0.330 |
| Fremantle Back Awareness Questionnaire | 0.00 [0.00, 2.50] | 6.00 [1.00, 15.00] | 0.00 [0.00, 1.00] | 5.00 [3.00, 7.00] | 1.00 [0.00, 5.00] | 7.00 [3.00, 13.00] | 0.093 |
| Tampa scale of kinesiophobia | 34.00 [29.00, 39.00] | 34.00 [29.00, 38.00] | 36.00 [33.50, 41.00] | 30.50 [26.50, 35.80] | 34.00 [31.30, 39.00] | 31.00 [29.00, 34.00] | 0.273 |
| Spielberger's State Anxiety Inventory | 32.00 [26.50, 37.00] | 33.00 [27.00, 34.00] | 32.00 [26.50, 35.50] | 30.00 [26.30, 33.50] | 31.00 [28.00, 35.00] | 34.00 [31.00, 38.00] | 0.433 |
| Spielberger's Trait Anxiety Inventory | 38.00 [35.00, 49.00] | 43.00 [38.00, 50.00] | 41.00 [39.00, 43.50] | 39.00 [34.50, 46.00] | 40.00 [35.80, 47.30] | 43.00 [38.00, 48.00] | 0.895 |
| Arbeitsindex Baecke | 2.14 [1.57, 3.43] | 2.14 [1.57, 3.71] | 1.86 [1.50, 2.07] | 1.57 [1.43, 1.96] | 1.71 [1.43, 2.25] | 1.86 [1.71, 2.14] | 0.013 |
| Sports score Baecke | 3.22 [2.21, 6.28] | 3.48 [1.37, 5.22] | 3.15 [2.60, 4.74] | 4.06 [2.71, 7.20] | 4.06 [2.90, 7.07] | 2.95 [0.126, 5.31] | 0.784 |
| Sportsindex Baecke | 3.25 [2.88, 3.63] | 3.00 [2.50, 3.75] | 3.00 [2.75, 3.25] | 3.25 [2.56, 3.75] | 3.25 [3.00, 3.81] | 2.75 [2.25, 3.50] | 0.683 |
| Sleep quality | 7.00 [4.50, 8.00] | 6.00 [5.00, 8.00] | 6.00 [4.50, 7.00] | 5.00 [3.00, 7.00] | 6.00 [5.00, 7.00] | 6.00 [4.00, 7.00] | 0.197 |
| Expectation balance | 6.00 [4.00, 6.00] | 3.00 [1.00, 5.25] | 5.00 [3.00, 6.00] | 5.00 [3.00, 6.00] | 4.50 [3.00, 6.00] | 6.00 [3.00, 6.00] | 0.769 |
| Expectation pain | 7.00 [5.00, 8.00] | 6.00 [4.00, 6.00] | 7.00 [6.00, 8.00] | 6.00 [6.00, 7.00] | 7.00 [5.00, 8.00] | 6.00 [5.00, 7.00] | 0.591 |
| Oswestry disability index | NA | 26.00 [24.00, 30.00] | NA | 25.00 [22.50, 31.00] | NA | 24.00 [24.00, 28.00] | 0.866 |
| painDetect questionnaire | NA | 7.00 [4.00, 8.00] | NA | 6.00 [4.00, 9.75] | NA | 7.00 [5.00, 10.00] | 0.690 |
| Baseline Pain | NA | 4.00 [2.00, 6.00] | NA | 2.00 [1.00, 4.75] | NA | 3.00 [1.00, 5.00] | 0.237 |
| N(%), median [interquartile range]. Statistical tests refer to potential differences across interventions |  |  |  |  |  |  |  |

### 3.4 Mixed model with PW ratio as dependent variable (primary outcome)

The model fit the data well, with a conditional R^2^ value showing that 62.6 % of the variance in the PW ratio was explained by the full model, considering fixed and random effects. The marginal R^2^ value indicated that the fixed effects alone explained 17.1 % of the variance. The fixed effects results indicated a significant effect of surface (stable vs. foam) (F[1,422] = 68.45, p < 0.001, stronger ankle-steered PW on a stable surface). Age (F[1,135] = 0.31, p = 0.580) and BMI (F[1,135] = 2.44, p = 0.121) did not significantly influence the outcome, while sex had a significant effect (F[1,135] = 7.08, p = 0.009, females had stronger lumbar-steered PW in general). Interestingly, there was no significant effect of group (healthy participants vs. patients) (F[1,135] = 0.178, p = 0.674). Furthermore, the interaction between timepoint and intervention was significant (F[2,422] = 4.13, p = 0.017), indicating that the difference before-after the intervention depended on the type of intervention (Table 2 and Figure 4). Parameter estimates showed that there was a significant effect of LMANIP on the difference of the PW ratio before-after compared to the NI condition (β=-0.047, t(422)=-2.71 p=0.007), as well as compared to the LMOB condition (β=-0.039, t(422)=-2.17, p=0.030). There was no significant difference in the PW ratio difference before-after between LMOB and NI (β=0.008, t(422)=-0.475, p=0.635).

**Figure 4.**
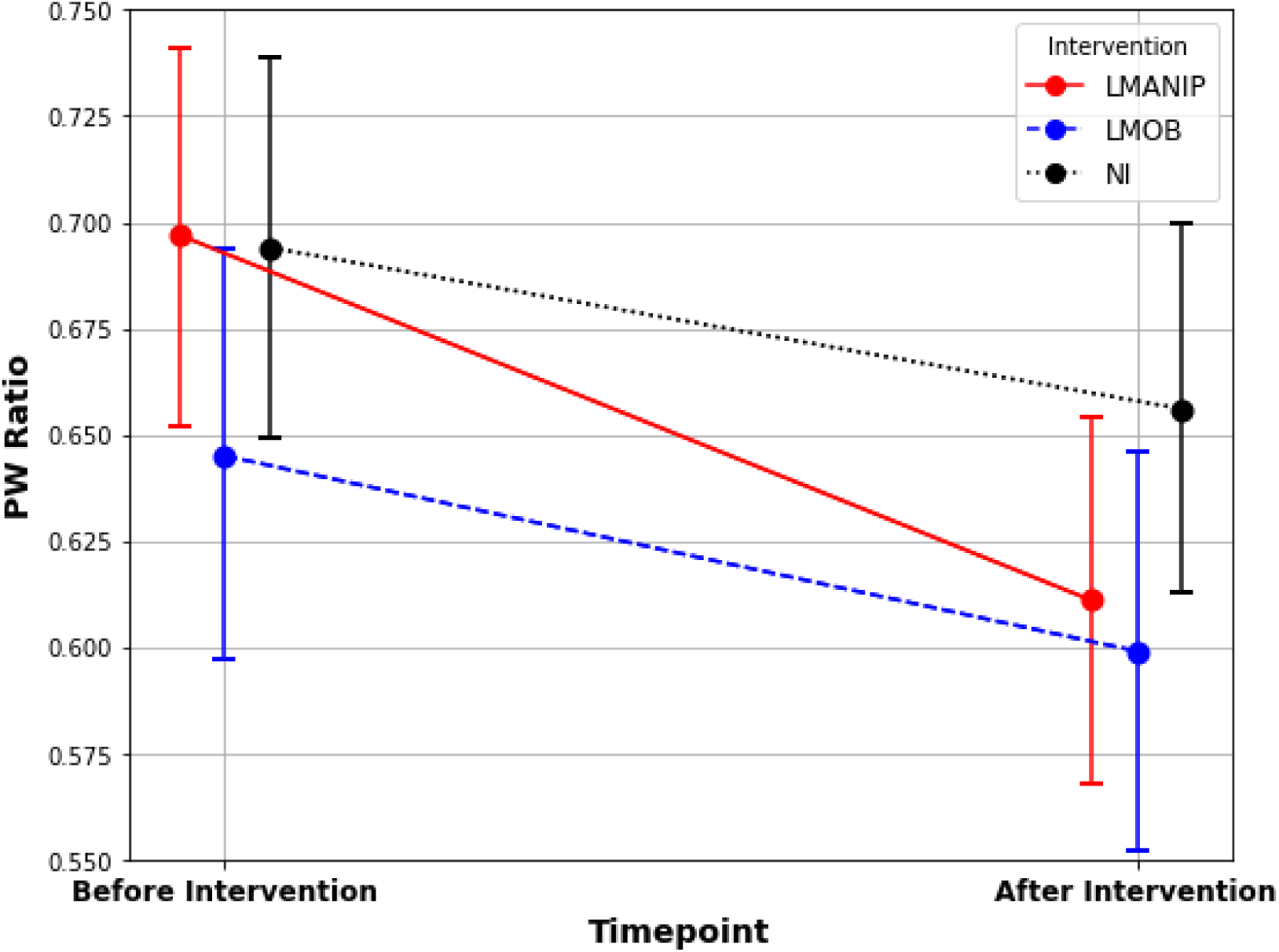
Proprioceptive weighting (PW) ratios across intervention and timepoints. PW ratios with 95 %CI (error bars) for the three interventions across timepoints. PW, proprioceptive weighting ratio; LMANIP, lumbar spinal manipulation; LMOB, lumbar spinal mobilization; NI, no intervention.

### 3.5 Mixed model with pain intensity (NPRS score) as dependent variable

A conditional R^2^ value of f 79.8 % indicated that the model generally fit the data very well. Fixed factors accounted for 29.9 % of the variance. The fixed-effects results showed significant effects of sex (F[1,43] = 9.24, p = 0.004), indicating that being female was generally associated with increased pain intensity ratings. BMI (F[1,43] = 0.12, p = 0.725) and age (F[1,43] = 2.59, p = 0.115) did not significantly influence the outcome. A significant 3-way interaction between timepoint, intervention and baseline PW ratio was observed (F[2,150] = 3.150, p = 0.017), suggesting that the effect of intervention on pain intensity ratings (before vs. after) varied based on the level of the baseline PW ratio and timepoint. With regard to this, the parameter estimates indicated that the (overall greater) effect of LMANIP on pain intensity changes (before vs. after) was moderated by the baseline PW ratio, with the difference in pain intensity change between LMANIP and LMOB being more pronounced with lower baseline PW ratio (β = -7.39, t(149) = -2.88, p = 0.004). Specifically, the negative β value suggested that lower baseline PW ratios were associated with greater pain reduction following LMANIP than following LMOB. Such a moderating effect of the baseline PW ratio was not observed for LMANIP compared to NI (β = -3.82, t(149) = -1.46, p = 0.145), nor for LMOB compared to NI (β = 3.57, t(149) = 1.49, p = 0.138) (Figure 5).

**Figure 5.**
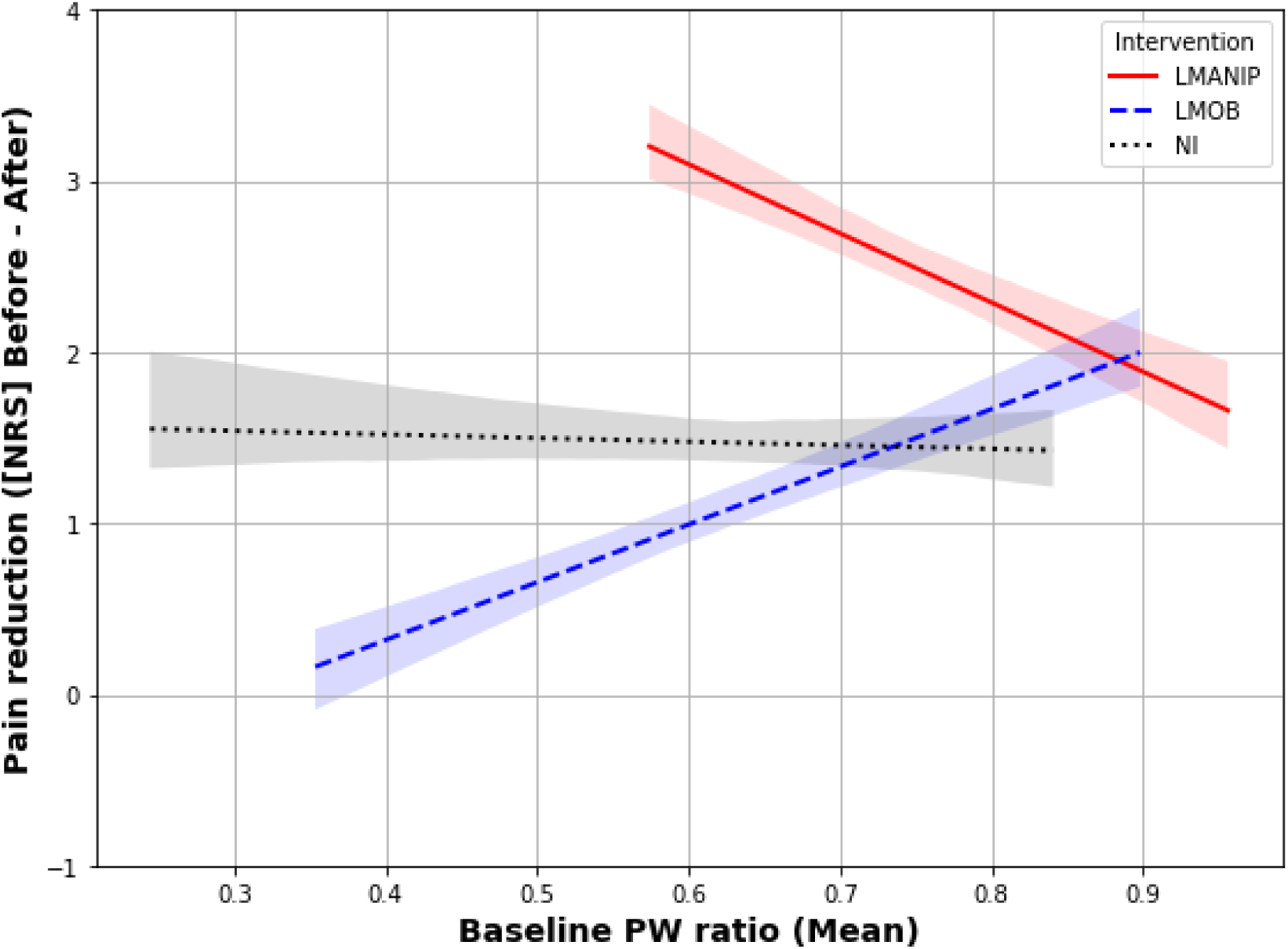
Pain reduction across interventions and its dependence on the baseline PW ratio. Y-axis: Pain reduction calculated based on pain ratings before-after (NPRS scores) for the three interventions (N = 52). Baseline PW, baseline proprioceptive weighting ratio averaged across surfaces (foam/stable); LMANIP, lumbar spinal manipulation; LMOB, lumbar spinal mobilization; NI, no intervention

### 3.6 Blinding

A chi-square goodness-of-fit test was conducted to assess whether the research assistants performing the data collection correctly guessed the intervention the participants received (LMANIP, LMOB, or NI). The test was not statistically significant, Χ² (4, N = 142) = 0.01, p = 0.906, indicating that the research assistants’ guesses were independent of the actual intervention received. A second chi-square test was performed to examine whether the clinician providing the intervention could identify whether the participant belonged to the patient or the healthy group. The results were significant, Χ² (1, N = 142) = 8.14, p = 0.004, suggesting that the clinician was able to distinguish between patients and healthy participants, even if there was no interaction other than providing the intervention involved. The contingency tables are reported in the supplementary material (Table S1).

## 4. Discussion

This single-blind randomized controlled trial investigated the effects of lumbar spinal manipulation, lumbar spinal mobilization, and no intervention on proprioceptive weighting (PW) in 142 adults with and without CLBP. The results indicate that a single SM session elicits an immediate (enhancing) effect on lumbar proprioceptive function irrespective of health status. Furthermore, SM was more effective in reducing clinical pain in patients with non-specific CLBP than the other two control conditions. However, this effect varied significantly depending on the baseline PW ratio, suggesting that the initial PW status may have predictive value for the analgesic response following SM.

Most theories underlying the clinical reasoning and explanations of the effects of SM interventions hypothesize a role for the nervous system [19,28,41]. However, there is a lack of well-controlled studies that provide evidence for this claim, and the relationship with potential analgesic effects remains unclear [29,58]. A common rationale for manual interventions is that spinal pain and dysfunction alter the processing of afferent feedback, potentially leading to disrupted sensorimotor integration [3]. Manual treatment is believed to help normalize this altered integration of afferent inputs, which can contribute to pain relief [20,48]. The current results provide solid evidence that lumbar SM has a notable impact on paraspinal proprioceptive mechanisms, potentially improving sensory integration in the lumbar spine, and enhancing balance and motor control. These findings are consistent with those of research indicating that manual therapy can positively affect sensorimotor processing and proprioception. Fryer and Pearce found a significant decrease in both corticospinal and spinal reflex excitability following SM, and suggested that these changes might alter motor recruitment strategies [15]. Clark et al. observed that SM led to a reduction in the stretch reflex of the erector spinae muscles. They proposed that SM might alter the activity of muscle spindles and other segmental sites involved in the Ia reflex pathway [8]. In contrast to these findings, other studies examining the effects of SM on postural sway and proprioception in patients with LBP have found minimal or no effects [17,30,44]. Goertz et al. found no change in postural sway in patients receiving manual therapy [17], while Learman et al. found no consistent effect of SM on trunk proprioception after SM compared to sham treatment [30]. Possible explanations for the non-consistent results could be that 1) postural sway is a complex readout that relies on afferent input from various body locations and tissues [17,21,30] and 2) measures of proprioceptive function such as joint repositioning sense can be confounded by motor skills or memory [51]. Vibrotactile stimulation at different key locations for balance control during standing and with eyes closed, as applied in the current study, allows to uncover the individual preferred proprioceptive source for balance control through evaluation of PW, which is a more direct assessment of lumbar proprioceptive function.

Interestingly, in this sample, no significant differences in PW ratios were observed between patients with CLBP and healthy participants (main effect “group”: F[1,135] = 0.21, p = 0.646/“group:surface” interaction F[1,421] = 1.59, p = 0.207), indicating similar PW strategies across groups. These findings are different from previous findings, in which patients with LBP seemed to adopt a body and trunk stiffening strategy and relied more on ankle proprioception to control their posture during quiet upright standing [4,5]. Here, age has been discussed as a potential explanation for the inconsistent findings regarding differential PW strategies between patients with LBP and healthy participants, because other studies found no differences in PW strategies in older populations with LBP [4,27]. However, the current study population was relatively young (mean: ∼26 years old), which might therefore not explain the non-significant differences in PW strategies between the groups in our sample. More research is needed to identify other possible factors influencing PW strategies, such as clinical pain at the time of testing, physical activity and sleep quality.

When interpreting changes in PW in isolation, it may seem plausible that pain reduction drives the observed shift in PW among those experiencing pain relief. However, the lack of a significant association between changes in PW and pain intensity (Spearman’s rho = 0.003, p = 0.969) raises doubts about a direct link between improved lumbar proprioceptive function and pain reduction, at least within a single-session design applied in this study. The significant three-way interaction between timepoint, intervention, and baseline PW ratio (F[2,150] = 3.150, p = 0.046) provides a more nuanced view of the relationship between PW and changes in pain intensity. While SM was generally more effective in reducing pain, it tended to result in greater pain reduction than spinal mobilization at lower baseline PW ratios, suggesting that SM may be more effective in patients with a stronger pre-intervention lumbar-steered PW. At first glance, this might seem counterintuitive, as SM has been shown to enhance lumbar-steered PW. The fact that patients with a stronger lumbar-steered PW profit more from SM suggests that these patients may be better equipped to respond to the mechanical input provided by SM. In these patients, the lumbar proprioceptive system may be in an optimal baseline state, allowing SM to enhance proprioceptive input and translate more directly into pain relief. These dynamic interactions between PW changes and pain relief highlight the need for future longitudinal studies to explore the long-term effects of SM on proprioception and pain outcomes. Understanding whether repeated SM sessions can amplify “proprioceptive benefits” and whether this translates into sustained pain relief could provide valuable insights for optimizing SM interventions.

This study has a few limitations. Only short-term outcomes were assessed, leaving the long-term effects of manual intervention on pain and proprioception unexplored. Moreover, the generalizability of the results is limited, as the study focused on a specific population with localized LBP without chronic pain in other body regions. This restricts the applicability of the findings to other subgroups such as patients with widespread pain. Another limitation was the clinician’s ability to distinguish between the groups, which could have influenced the intervention. Nevertheless, the participants were blinded to the study’s hypothesis, making any bias toward the PW ratio in a specific direction highly unlikely. Additionally, the lack of movement illusions resulted in the exclusion of a relatively high number of participants (N = 60). Most of these non-responders (∼88 %) reported no illusions during ML stimulation, potentially because of the differences in muscle architecture between ML and MTS. In parallel-fibred muscles, such as MTS, vibration may more effectively activate muscle spindles through tendon co-stimulation [49]. It is not uncommon for 10-20 % of participants to be non-responders regarding illusions during tendon co-stimulation (as it is the case during MTS stimulation), and they are typically excluded from analyses [9,42,49]. This exclusion is necessary because during the trial-and-error process to verify the vibrator’s ability to provoke movement illusions (which was not done in the current study), participants may become aware of the expected stimulation outcome. Even without explicit information about the illusory movement, this awareness can lead participants to imagine the expected movement, which can distort the results [49]. Finally, the initial sample size calculation was performed for a healthy control group, and a patient-specific calculation was not conducted. Although this limitation may affect the generalizability and robustness of the findings, adjustments were made to assess the robustness of our results in patients with the available sample size. We explored different random-effects structures by comparing the models with random intercepts and random slopes. Consistency in fixed effect estimates across these models suggested that results in were not highly sensitive to the assumptions regarding random effects. Additionally, we tested alternative covariance structures (compound symmetry and autoregressive AR(1)/ARMA(1,1)) to evaluate the robustness of the model further. Stable fixed-effect estimates across these covariance structures indicated that the results were unlikely to be affected by assumptions related to covariance.

In conclusion, the current findings provide the first evidence that lumbar proprioceptive input is ’up-weighted’ following SM, aligning with and extending findings from pre-clinical research on SM-like interventions in animal models [40]. Furthermore, the initial PW status may be a potential biomarker for predicting the efficacy of SM. Further research is necessary to fully understand the long-term implications and establish its predictive value for SM outcomes, aiding the development of personalized treatment strategies for patients with CLBP.

## Data Availability

All datasets analyzed in this study are available in the main text or upon request. Further information and requests should be directed to the corresponding author, Michael L Meier at the following e-mail address: and will be considered on a case-by-case basis, in accordance with the local policies of the sponsor and the relevant Swiss and European Union data protection and privacy legislation.

## 5. Acknowledgments

This project was funded by the European Centre for Chiropractic Research Excellence (ECCRE) and the Balgrist Foundation. The trial was preregistered at ClinicalTrials.gov (NCT 04869514) without an analysis plan. We thank the REVAL Research Center (Hasselt University) for providing and maintaining the materials used for muscle vibrations. Additionally, we thank Selina Schnider for helping us finalize the manuscript.

## Ethical approval

This study was approved by the Swiss local ethics board ‘Kantonale Ethikkommission Zürich, KEK (Nr. 2021-01331). All procedures adhered to the Declaration of Helsinki and current legislation pertaining to the management of personal data.

## Data availability statement

The authors have declared that no competing interests exist.

