## Supplementary material for "The Impact of Spinal Manipulation on Lumbar Proprioception and its Link to Pain Relief: A Randomized Controlled Trial": S1

**S1. Blinding results**

| Intervention received | Guess research assistants | | | | | | |
| --- | --- | --- | --- | --- | --- | --- | --- |
|  | ***LMANIP*** | | ***LMOB*** | | ***NI*** | | ***Total*** |
| *LMANIP* | 23 | | 19 | | 7 | | 49 |
| *LMOB* | 21 | | 9 | | 18 | | 48 |
| *NI* | 21 | | 20 | | 14 | | 45 |
| *Total* | 55 | | 48 | | 39 | | 142 |
| Chi-Square: 0.014, p = 0.906 | | | | | | | |
| Group | | **Guess chiropractor** | | | | | |
|  |  | ***HC*** | | ***PAT*** | | ***Total*** | |
| *HC* | | 65 | | 29 | | 94 | |
| *PAT* | | 25 | | 23 | | 48 | |
| *Total* | | 90 | | 52 | | 142 | |
| Chi-Square: 8.141, p = 0.004 | | | | | | | |

**S2. Linear mixed models with center of pressure (COP) displacements as dependent variable**

To test whether the experience of movement illusions at each location (MTS and ML) impacted postural sway (dependent variable: mean COP[mm]), two separate mixed models (for each location) were created. The models included fixed effects for "sex” (male/female), "age", "bmi, “group" (healthy control/patient), "timepoint" (before intervention/after intervention), "surface" (stable/foam), “illusion” (yes/no) and their interactions ("surface:illusion" and “timepoint:surface:illusion”). The interaction effects of "group:illusion" and “group:surface:illusion" were included in the model but were subsequently removed after being found non-significant, in order to focus on estimating the main effects more precisely.

The linear mixed model analysis for location ML (conditional R^2^ = 51.9%, marginal R^2^ = 11.5%) revealed a significant three-way interaction between timepoint, surface, and illusion (F[1,600] = 4.02, p = 0.045) indicating that the experience of a movement illusion during ML stimulation significantly affected COP displacements depending on the surface and timepoint. This interaction was mainly driven by a significant effect of illusion vs. no illusion on COP displacements before intervention on the foam surface (post hoc t-tests, t(470) = 2.07, p = 0.038, Figure S2). No significant effects were found for MTS location (surface:illusion, p = 0.058; timepoint, surface; illusion, p = 0.713). In addition, fixed effects analysis yielded a significant main effect for “sex” in the MTS model (F[1,196] = 27.451, p < 0.001 / men had generally stronger COP displacements) and a significant main effect of “bmi” in the ML model (F[1,196] = 14.51, p < 0.001 / the higher the bmi the lower the COP displacements).

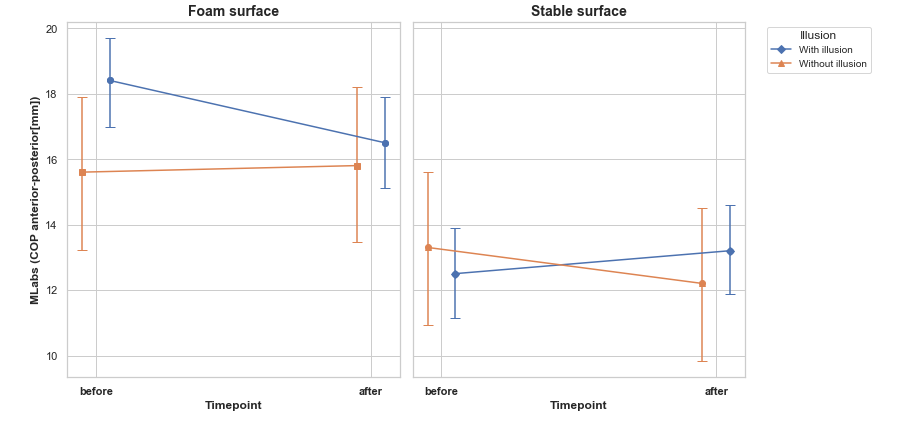

***Figure S2. Impact of movement illusions on center of pressure (COP) displacements during vibrotactile stimulation of the M. longissimus (ML).*** *COP changes with 95 %CI (error bars) for the two conditions (with/without illusion) and for each surface (stable/foam). MLabs represents the absolute displacement of the mean COP [mm] during M. longissimus vibration.*
